# Evaluating public and patient involvement in interventional research – a newly developed checklist (EPPIIC) with application to the COB-MS feasibility trial

**DOI:** 10.1101/2024.03.17.24304433

**Authors:** Elise Pyne, Robert Joyce, Christopher P. Dwyer, Sinéad M. Hynes

## Abstract

Public and patient involvement (PPI) has been identified as an increasingly desired and, often, required component of trial methodology – leading to higher quality, more accessible and relevant clinical research, alongside increased recruitment, funding success and insight into research impact. However, despite the great variety of frameworks and checklists available for assessing PPI, most are limited with respect to important features (e.g. applicable in specific contexts only, fail to clarify what should be assessed and reported, lack the necessary comprehensiveness or are biased in favour of researcher reporting). Thus, the current research aimed to address such limitations through the development of a new checklist, the EPPIIC, through review, thematic analysis and ‘meta-evaluation’ in conjunction with PPI engagement. A further aim was to pilot the EPPIIC through its application to and reporting on the COB-MS trial, which utilised PPI throughout the research’s life-cycle. Upon completion of the EPPIIC, three thematic ‘sub-scales’ emerged: (1) Policy & Practice, (2) Participatory Culture and (3) Influence & Impact. All findings are presented and discussed in light of theory and research. Notably, findings recommend EPPIIC as a useful means of assessing PPI in future trials.

## Introduction

Public and patient involvement (PPI) is vital for trial methodology, leading to higher quality, more accessible and relevant clinical research (1). It is also rapidly becoming a more-and-more desired – if not required – component of clinical trials (2). Recent approaches to PPI aim to empower and enable such member involvement, allowing for flexible structures and procedures created by both PPI members and researchers. ‘*Nothing about us without us’* – a message often used by PPI members in context – tells clinical researchers that the raw purpose of their work is to improve the lives of those affected by the topic of their study (3). While researchers may understand the intricate pathology of disease, it is patients who have the unique lived experience of the condition.

PPI inclusion further increases recruitment, funding success rates and provides unique insights into the potential impacts of the research (4). For PPI members, involvement can increase skills and boost feelings of self-worth and confidence (5). Notably, *true* PPI extends beyond mere consultation to active partnership throughout the research’s life-cycle – from funding application and protocol development all the way to dissemination and knowledge translation (6). Indeed, through appropriate implementation, PPI members can be the invaluable ‘critical friends’ needed to improve the overall quality of clinical research (7–8).

With pressure from funders to embed PPI into clinical research and improved awareness of the benefits of PPI, the rate of PPI in clinical research has rapidly increased (2;9). However, the evidence of PPI impact is less clear, with continued discussion and debate concerning the means of evaluating the use of PPI in clinical trials (10–11). Though there is vast agreement regarding the need to capture the negative and positive aspects of PPI processes (10), there exists a variety of frameworks, surveys and checklists – with diverse perspectives that claim to capture the challenges faced and opportunities created when using PPI in clinical research (12–13). However, the comprehensiveness and focuses of these tools are debatable in light of this diverse pool from which to choose – debate further reinforced by the relative recency of PPI as a phenomenon in clinical trial methodology. Not surprisingly, there also exists a demand for a guideline and/or framework that not only evaluates PPI, but also provides researchers with clarity regarding what *should be* assessed and reported, in context (12). Again, ‘context is key’ and, unfortunately, not all PPI evaluation strategies are contextually appropriate, when such comprehensiveness is desired. Indeed, comparison and appraisal of strategies, assessing impact and ensuring what is claimed has been done, are at the heart of evaluating PPI approaches.

None of these extant PPI checklists are without their limitations (13). Most frameworks evaluate from the researcher’s perspective – a strategy that immediately suggests reporting bias. On the other hand, evaluation strategies that do account for PPI members responding are also problematic, with one review finding that only 11.1% of tools had the reading level sufficient for public or lay persons’ understanding (11). Many of these also fail to address the same areas from both perspectives-the PPI member(s) and the researcher(s). Typically PPI members are only questioned on their input rather than the accommodations that have been made for them by the researcher team, whereas researchers have the opportunity to comment on both.

Thus, the focus of the current research is to address the limitations of previous evaluation tools through the development of a new checklist, that is generalisable across research typologies within the parameters of clinical interventions through comprehensive description of PPI focuses, though non-specific reporting cues (e.g. with added focus on open-ended reporting). This has been achieved through two overarching aims and through the following objectives:

Aim 1: Develop a checklist that can be applied across a variety of research settings:

1. To conduct a review of current PPI guidelines and outcome measures.
2. To assess the identified outcome measures and collate thematically identified focuses to create an evaluation checklist (or checklists) to appraise the quality of PPI within trials.

Aim 2: To pilot this newly developed checklist:

1. 3. For application to a trial, Cognitive Occupation-Based programme for people with MS (COB-MS; 14-15; described further in the methods section), to demonstrate its use; and
2. 4. To report the COB-MS PPI evaluation.

## Materials and Methods

To note-the COB-MS PPI member (RJ) was involved throughout the research process described here (and the COB-MS trial) and had a lead role in directing the research.

## Aim 1 Methods

### Review of current tools

A literature review of extant PPI checklists was conducted, focusing on the process and outcome assessments of PPI. These checklists were then subjected to content analysis to identify common topics throughout and identify the exact quantitative and qualitative questions and methods to evaluate PPI. This work was completed in consultation with the COB-MS PPI member (RJ). This was carried out with the overall aim of assessing PPI in the COB-MS feasibility trial (14–15). Variations in the style of existing checklists also provided insight in the best way to formulate questions.

### Search Strategy

To navigate and formulate the research question, the SPIDER search strategy was used (16; see Table 1) to ensure the relevant and appropriate frameworks were being evaluated. Other search methods included a keywords search using PubMed and Google Scholar search engines. Boolean operators were used to carry out an extensive search of all related research and documentation (see Table 2).

**Table 1:** Identification of research question through SPIDER Search strategy.

| **S**<br>**Sample** | **PI**<br>**Phenomenon of Interest** | **D**<br>**Design** | **E**<br>**Evaluation** | **R**<br>**Research**<br>**Type** |
| --- | --- | --- | --- | --- |
| Various PPI evaluation frameworks used in clinical trials | Efficacy of framework to report PPI | Using PPI Evaluation frameworks and modifying them to fit this trial | Impact of PPI on study recruitment, retention, and overall trial quality | Mixed methods |

**Table 2:** Comprehensive list of keywords used in Boolean operators to identify research concerning PPI evaluation strategies.

|  |  |  |  |  |
| --- | --- | --- | --- | --- |
| Public and Patient Involvement OR PPI OR Patient Engagement OR Patient Participation OR User Involvement | AND | Framework OR checklist OR criteria OR agenda OR Method OR Approach OR Guideline OR model OR Toolkit OR Strategy | AND | Evaluation OR Assessment OR Appraisal |

In addition to the literature review, the Centre of Excellence for Partnership with Patients and the Public (CEPPP) database was also used to identify relevant checklists for this research. The CEPPP is an online resource that encompasses a number of evaluation tools, to enable researchers to assess the quality of PPI in their research. The CEPPP evaluates the included tools based on usability, comprehensiveness, patient and public perspective and scientific rigour by applying targeted questions to each framework. From CEPPP’s previous evaluation, each framework was investigated, with 11 satisfying our inclusion criteria, included in Table 3 below.

**Table 3:** Summary of themes and subthemes found through a literature review of PPI evaluation tools.

| <u>Theme and Subtheme</u> | <u>Checklists</u> |
| --- | --- |
| **Policy & Practice** |  |
| Planned Strategy and Methods | (13; 19- 34), |
| Resource mobilisation | (13; 19-20; 22-28; 31; 33; 35-37) |
| Reports of PPI | (13; 19- 24; 26; 28; 31; 33-38) |
| Recruitment | (32; 35) |
| Team Engagement | (21-25; 28;31-32) |
| Adaptability | (13; 19-20; 22; 28; 31; 36-37) |
| Experience and representation | (13; 22-25; 27; 31; 35) |
| Management, and Implementation of PPI Recommendations | (13; 19-22; 28; 38) |
| Communication methods | (23-24; 27; 31-32; 34) |
| **Participatory Culture / Collaboration** |  |
| Boosting Awareness | (19-20; 23-24; 28) |
| Participatory Feedback | (19-20; 22-24; 26; 31; 33-34) |
| **Influencing Outcomes of PPI** | (13; 21; 35) |

### Inclusion / exclusion criteria

The only inclusion criterion for framework evaluation was that the framework must assess PPI in the context of clinical research. Frameworks were excluded if they were not relevant to research or deviated from it -e.g. solely focused on team dynamics and collaboration.

### Analysis of Extant Frameworks

After all relevant checklists had been identified, thematic analysis was conducted, to identify, analyse and report themes within the qualitative data. Specifically, data were analysed consistent with Braun and Clarke’s (17–18) six-phase analytic process, which highlights three main tasks: familiarisation with data; coding and theme identification; and the reviewing and refining of themes. This method included reading and re-reading of frameworks to a gain familiarity with the materials, prior to identification of components of interesting elements, codes, and approaches. An extensive list was generated and sorted into overarching themes.

From this list of themes, a content analysis was completed to formulate the quantitative and qualitative questions required for a comprehensive checklist. Approaches to PPI were divided into formulative and summative questioning, meaning that while some questions related to the process of PPI, others dealt with the final outcomes instead. Developed questions were reviewed to remove any questions that were overlapping, repetitive or were not directly relevant to PPI. Reviewing and refining the checklist items until finalisation and application of the checklist involved the PPI members, research team and an independent researcher who had no involvement with the COB-MS trial under assessment (i.e. to limit potential for bias).

## Aim 2 Methods

### Applying the newly developed checklist

Following its completion, the checklist was actively applied to evaluate the PPI associated with the COB-MS trial, ISRCTN11462710, (see 14-15), which utilised PPI throughout the trial’s life-cycle. Specifically, the trial partnered a PPI member as a contracted researcher [becoming an *embedded patient researcher* (EPR; 7)], included two PPI members in the Trial Steering Committee and created an external PPI consultation group.

## Results

### Aim 1 Results

Through thematic analysis, many overlapping patterns of themes were found. Subthemes were grouped into their overarching theme to create each of the three main themes. A summary of the research evidence that led to these subthemes is in the Table 3 below.

Three main themes were identified: (1) Policy & Practice, (2) Participatory Culture, and (3) Influence & Impact. *Policy & Practice* focused on the structure and strategy of PPI implementation prior to the research starting, centring on the efficiency and methodological aspects of PPI in research – the justification for which was to assess the preparedness of an organisation to carry out this type of research. This item was included as a measure of meaningful involvement and appropriate consideration of PPI. *Participatory Culture* referred to consideration of factors that could enhance or hinder PPI throughout the project, with focus on the research team’s ability to accommodate PPI and how PPI can be optimally engaged/integrated. Notably, this theme is perhaps the main discussion point for comparing trials that include PPI, focusing on the needs of the PPI members and how to appropriately involve them. Finally, *Influence & Impact* focused largely on outcomes of PPI, ensuring that the experience was beneficial to both researchers and PPI members. It reflects the overall impact of PPI on the research question and exhibits whether the PPI strategies initially proposed were actually implemented. This theme is of particular importance, as it not only reveals the advantage(s) of PPI within the trial being evaluated, but also has the capacity to facilitate recommendations for PPI within future interventions.

Following thematic analysis, relevant questions from extant checklists were collated under their relevant themes and subthemes (see Table 3). After filtering initial questions (e.g. by relevance, overlap and repetition) and amending as appropriate, the resulting questions were organised into two separate forms: researcher evaluation and PPI member evaluation. The final questions were generated by reviewing the wording of questions within other checklists and analysing clarity and comprehensiveness. Some of these were used verbatim, while others were adjusted to ensure better understanding of the question. This process was completed in close collaboration with the PPI member (RJ). Such organisation reduces bias and facilitates the ability of independent adjudicators to observe perspectives from distinct sources and compare them. Checklist items were tailored to each audience (e.g. with respect to ensuring accessible language) but were otherwise commensurate; though the researcher form includes some additional items (e.g. related to the overall trial budget). Items in the checklists were presented through means of both ‘box-ticking’ (i.e. both Likert scale and dichotomous, yes/no responding, as appropriate) and open-ended response. A balanced mix of formative/summative and qualitative/quantitative inquests were included in both forms.

The finalised checklists, titled “Evaluation of PPI for Interventional research Checklist (EPPIIC)” can be found in Supporting Information 1 and 2 (S1 and S2 Appendices). A summary of the steps taken in developing and applying the EPPIIC are in Table 4.

**Table 4:** Summary of steps involved in creation of new EPPIIC.

|  | Step | Resources/People involved | Results |
| --- | --- | --- | --- |
| 1 | Compile full list of available checklists | CEPPP Database search (PubMed, Google Scholar) | Total checklists identified (n= 20; 11 CEPP and nine database search) |
| 2 | Generate full list of themes | All identified checklists; PPI involvement | Total number of items in the first iteration of the checklist = 34 |
| 3 | Matching each item with a checklist item | All identified checklists | Checklist items were sorted into each identified theme |
| 4 | Grouping subthemes into relevant main themes | PPI involvement; research team discussion | Three overarching themes identified |
| 5 | Refining the items | PPI involvement; research team discussion | Decision to separate PPI and researcher evaluation |
| 6 | Piloting of the checklist | PPI and research team | Adjustments between PPI member and researcher surveys – e.g. change language, merging of items etc. |
| 7 | Finalising the checklist | PPI and research team | Number of items – ensuring equal questions between researcher and PPI member survey with appropriate balance of qualitative and quantitative questioning. |
| 8 | Application of the checklist | EPPIIC applied to the COB-MS trial | Results presented below. |

### Aim 2 Results

The EPPIIC (PPI member form) was completed by the trial’s EPR, RJ, while the researcher form was completed by trial PI, SMH. Overall, the checklists identified areas of strengths in terms of PPI inclusion and highlighted areas for future improvement. The results of the application of the checklist are presented under the three themes of the EPPIICs. Direct quotes from the checklists are presented for further context. These results are summarised in Tables 5, 6 and 7, with further description of presented in-text below. Notably, these tables focus on the PPI Member/EPR and researcher perspectives and translates a ‘tick-box’ exercise into meaningful results. The checklist asked both cohorts to provide their opinions to statements through a Likert ‘significance’ or ‘agreement’ scale. The box that was chosen by the respondent reflected the strength of their opinion towards the statement provided in each section.

**Table 5:**
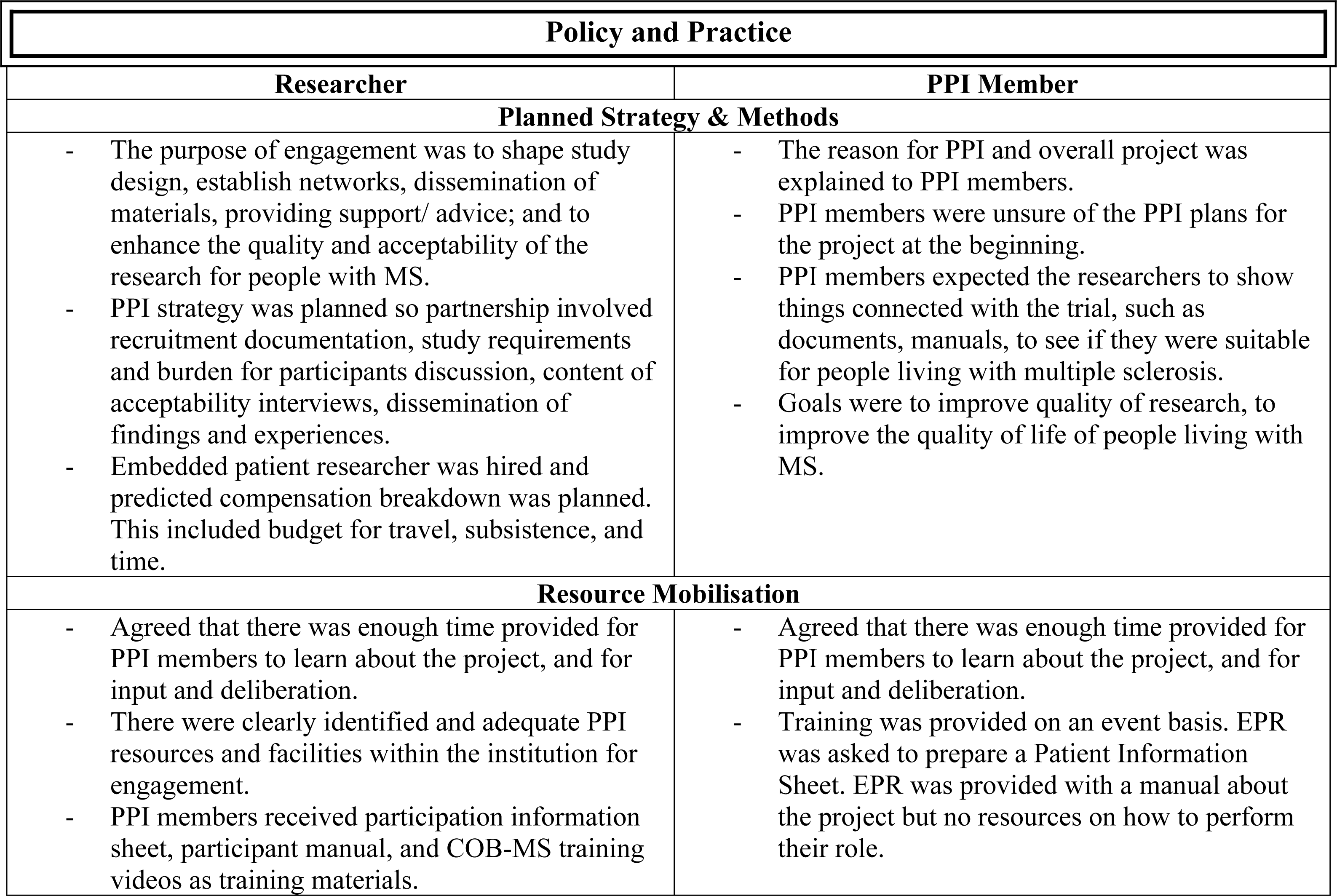
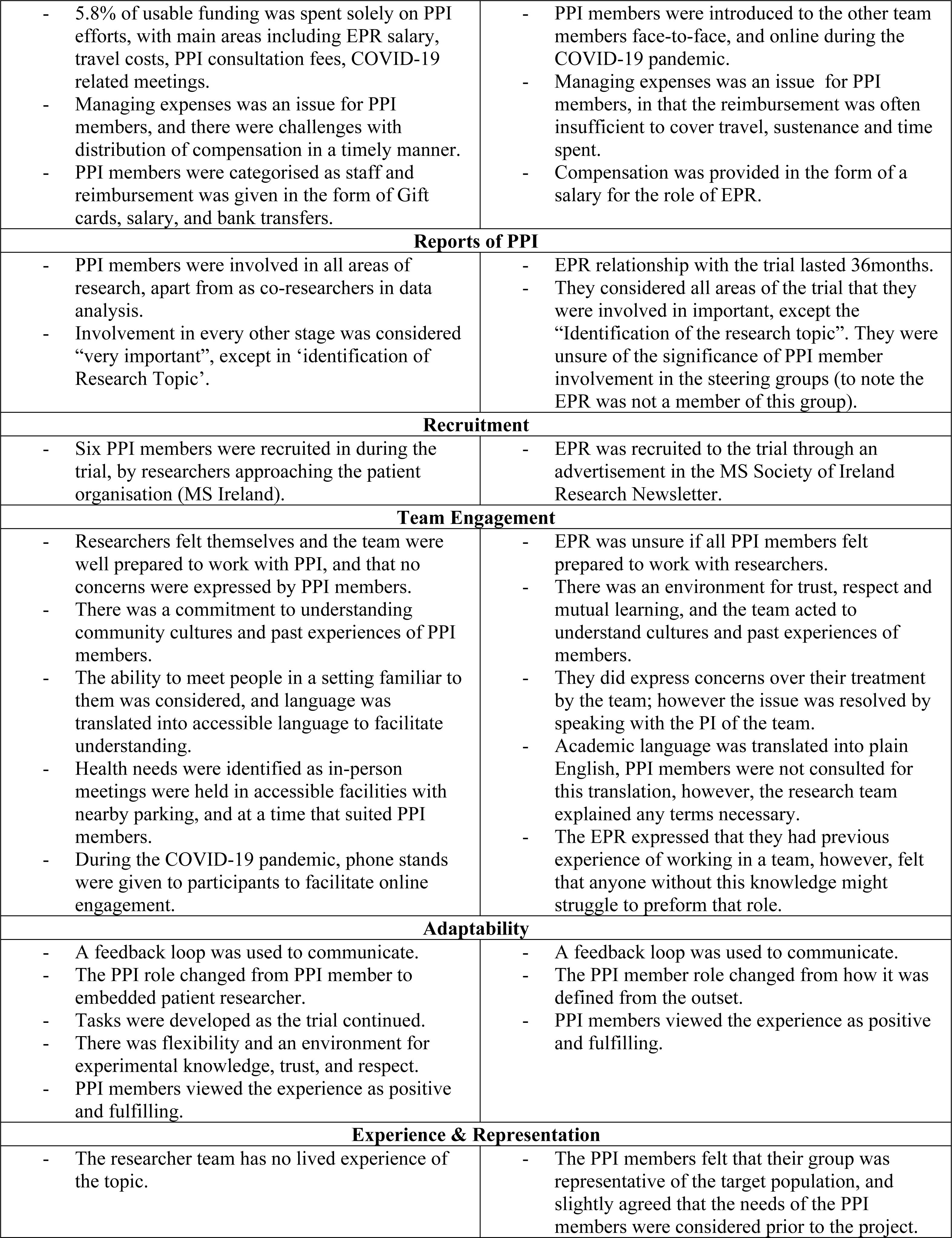
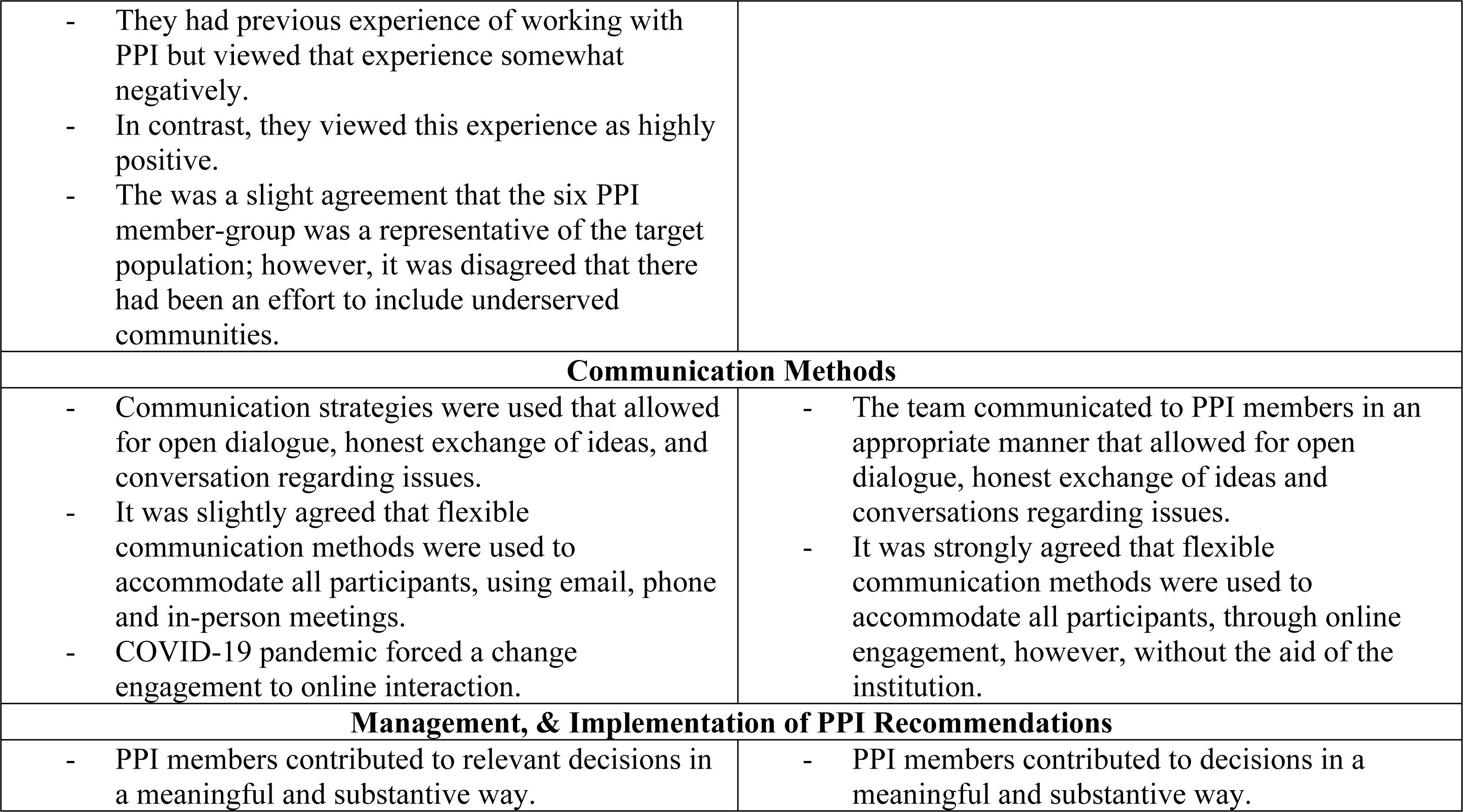
Summary of results from completed Researcher, and PPI Member EPPIIC in the Policy and Practice theme.

| Policy and Practice |  |
| --- | --- |
| Researcher | PPI Member |
| Planned Strategy & Methods |  |
| - The purpose of engagement was to shape study design, establish networks, dissemination of materials, providing support/ advice; and to enhance the quality and acceptability of the research for people with MS.<br>- PPI strategy was planned so partnership involved recruitment documentation, study requirements and burden for participants discussion, content of acceptability interviews, dissemination of findings and experiences.<br>- Embedded patient researcher was hired and predicted compensation breakdown was planned. This included budget for travel, subsistence, and time. | - The reason for PPI and overall project was explained to PPI members.<br>- PPI members were unsure of the PPI plans for the project at the beginning.<br>- PPI members expected the researchers to show things connected with the trial, such as documents, manuals, to see if they were suitable for people living with multiple sclerosis.<br>- Goals were to improve quality of research, to improve the quality of life of people living with MS. |
| Resource Mobilisation |  |
| - Agreed that there was enough time provided for PPI members to learn about the project, and for input and deliberation.<br>- There were clearly identified and adequate PPI resources and facilities within the institution for engagement.<br>- PPI members received participation information sheet, participant manual, and COB-MS training videos as training materials. | - Agreed that there was enough time provided for PPI members to learn about the project, and for input and deliberation.<br>- Training was provided on an event basis. EPR was asked to prepare a Patient Information Sheet. EPR was provided with a manual about the project but no resources on how to perform their role. |
| - 5.8% of usable funding was spent solely on PPI efforts, with main areas including EPR salary, travel costs, PPI consultation fees, COVID-19 related meetings.<br>- Managing expenses was an issue for PPI members, and there were challenges with distribution of compensation in a timely manner.<br>- PPI members were categorised as staff and reimbursement was given in the form of Gift cards, salary, and bank transfers. | - PPI members were introduced to the other team members face-to-face, and online during the COVID-19 pandemic.<br>- Managing expenses was an issue for PPI members, in that the reimbursement was often insufficient to cover travel, sustenance and time spent.<br>- Compensation was provided in the form of a salary for the role of EPR. |
| **Reports of PPI** |  |
| - PPI members were involved in all areas of research, apart from as co-researchers in data analysis.<br>- Involvement in every other stage was considered “very important”, except in ‘identification of Research Topic’. | - EPR relationship with the trial lasted 36 months.<br>- They considered all areas of the trial that they were involved in important, except the “Identification of the research topic”. They were unsure of the significance of PPI member involvement in the steering groups (to note the EPR was not a member of this group). |
| **Recruitment** |  |
| - Six PPI members were recruited in during the trial, by researchers approaching the patient organisation (MS Ireland). | - EPR was recruited to the trial through an advertisement in the MS Society of Ireland Research Newsletter. |
| **Team Engagement** |  |
| - Researchers felt themselves and the team were well prepared to work with PPI, and that no concerns were expressed by PPI members.<br>- There was a commitment to understanding community cultures and past experiences of PPI members.<br>- The ability to meet people in a setting familiar to them was considered, and language was translated into accessible language to facilitate understanding.<br>- Health needs were identified as in-person meetings were held in accessible facilities with nearby parking, and at a time that suited PPI members.<br>- During the COVID-19 pandemic, phone stands were given to participants to facilitate online engagement. | - EPR was unsure if all PPI members felt prepared to work with researchers.<br>- There was an environment for trust, respect and mutual learning, and the team acted to understand cultures and past experiences of members.<br>- They did express concerns over their treatment by the team; however the issue was resolved by speaking with the PI of the team.<br>- Academic language was translated into plain English, PPI members were not consulted for this translation, however, the research team explained any terms necessary.<br>- The EPR expressed that they had previous experience of working in a team, however, felt that anyone without this knowledge might struggle to perform that role. |
| **Adaptability** |  |
| - A feedback loop was used to communicate.<br>- The PPI role changed from PPI member to embedded patient researcher.<br>- Tasks were developed as the trial continued.<br>- There was flexibility and an environment for experimental knowledge, trust, and respect.<br>- PPI members viewed the experience as positive and fulfilling. | - A feedback loop was used to communicate.<br>- The PPI member role changed from how it was defined from the outset.<br>- PPI members viewed the experience as positive and fulfilling. |
| **Experience & Representation** |  |
| - The researcher team has no lived experience of the topic. | - The PPI members felt that their group was representative of the target population, and slightly agreed that the needs of the PPI members were considered prior to the project. |
| - They had previous experience of working with PPI but viewed that experience somewhat negatively.<br>- In contrast, they viewed this experience as highly positive.<br>- There was a slight agreement that the six PPI member-group was a representative of the target population; however, it was disagreed that there had been an effort to include underserved communities. |  |
| **Communication Methods** |  |
| - Communication strategies were used that allowed for open dialogue, honest exchange of ideas, and conversation regarding issues.<br>- It was slightly agreed that flexible communication methods were used to accommodate all participants, using email, phone and in-person meetings.<br>- COVID-19 pandemic forced a change engagement to online interaction. | - The team communicated to PPI members in an appropriate manner that allowed for open dialogue, honest exchange of ideas and conversations regarding issues.<br>- It was strongly agreed that flexible communication methods were used to accommodate all participants, through online engagement, however, without the aid of the institution. |
| **Management, & Implementation of PPI Recommendations** |  |
| - PPI members contributed to relevant decisions in a meaningful and substantive way. | - PPI members contributed to decisions in a meaningful and substantive way. |

**Table 6:**
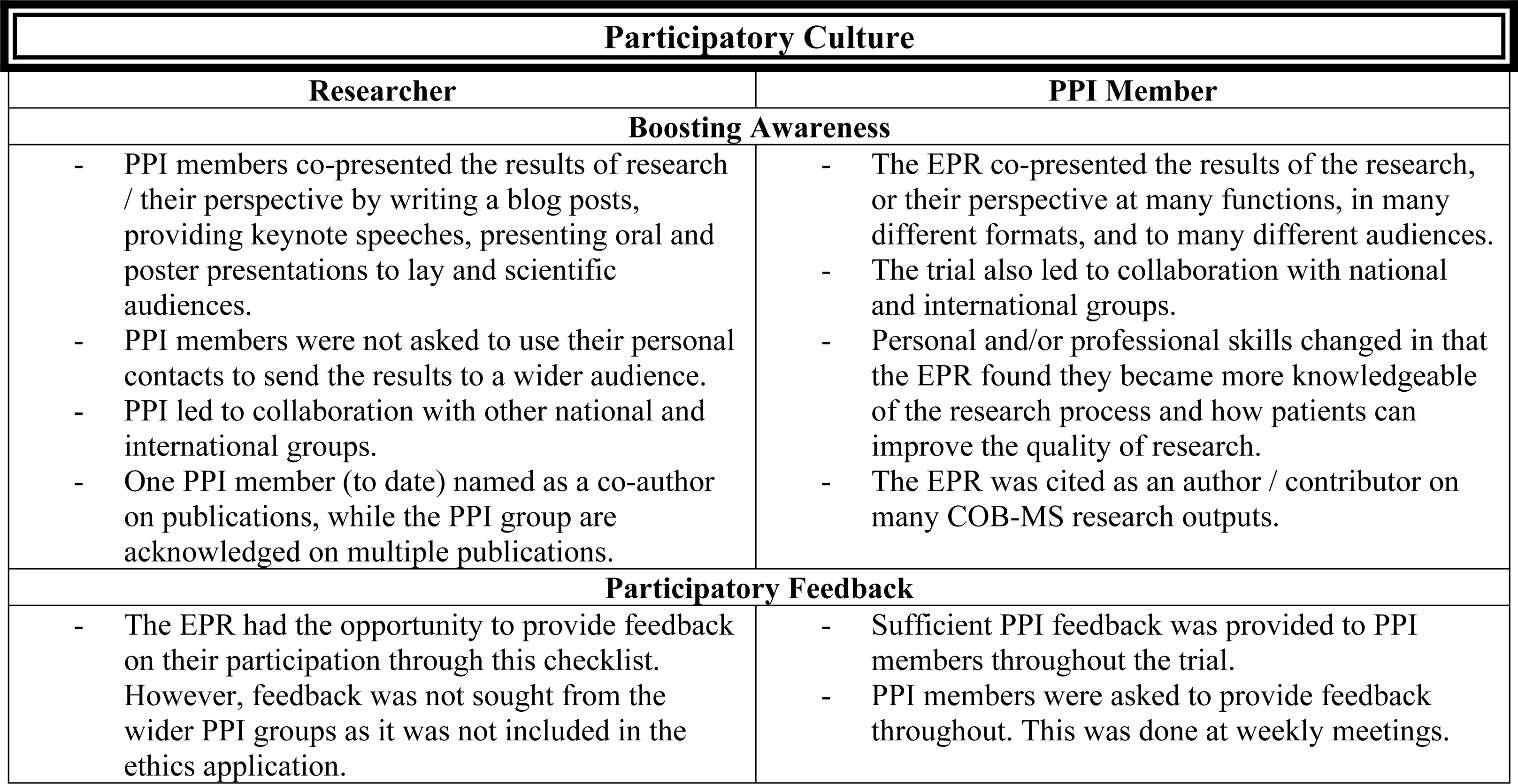
Summary of results from Researcher and PPI Member EPPIIC under the theme of Participatory Culture.

| Participatory Culture |  |
| --- | --- |
| Researcher | PPI Member |
| Boosting Awareness |  |
| - PPI members co-presented the results of research / their perspective by writing a blog posts, providing keynote speeches, presenting oral and poster presentations to lay and scientific audiences.<br>- PPI members were not asked to use their personal contacts to send the results to a wider audience.<br>- PPI led to collaboration with other national and international groups.<br>- One PPI member (to date) named as a co-author on publications, while the PPI group are acknowledged on multiple publications. | - The EPR co-presented the results of the research, or their perspective at many functions, in many different formats, and to many different audiences.<br>- The trial also led to collaboration with national and international groups.<br>- Personal and/or professional skills changed in that the EPR found they became more knowledgeable of the research process and how patients can improve the quality of research.<br>- The EPR was cited as an author / contributor on many COB-MS research outputs. |
| Participatory Feedback |  |
| - The EPR had the opportunity to provide feedback on their participation through this checklist. However, feedback was not sought from the wider PPI groups as it was not included in the ethics application. | - Sufficient PPI feedback was provided to PPI members throughout the trial.<br>- PPI members were asked to provide feedback throughout. This was done at weekly meetings. |

**Table 7:** Summary of notes and ideas in Researcher and PPI Member EPPIIC under the theme of Influencing the Outcomes of PPI.

| Influencing the Outcomes of PPI |  |
| --- | --- |
| Researcher | PPI Member |
| - Challenges of PPI in this trial included inclusion of PPI members when meetings were not chaired by the research team, and retention of PPI members during the COVID-19 pandemic due to unfamiliarity with Zoom/online meetings.<br>- The university’s process surrounding payment hindered the reputation of PPI in the trial.<br>- Motivation from the PPI panel and EPR enabled the impact of PPI.<br>- Having an EPR made the wider PPI tasks more successful and easier to manage.<br>- The term ‘*Embedded Patient Researcher*’ was coined.<br>- The researcher would be likely to involve PPI in future research.<br>- Positive impacts on PPI in research included recruitment, developing patient materials, integration, engagement with trial participants, communication & dissemination plan, and collaboration. | - Challenges of PPI inclusion: 1) the university facilities were often not appropriate for the PPI members; and 2) did not provide any equipment resources.<br>- PPI member quality of life changed as their involvement allowed them to create a positive outcome from a negative situation and helped them find purpose.<br>- In the future, a buddy system could be proposed to involve inexperienced PPI members.<br>- Currently the social welfare system in Ireland does not permit people in receipt of some payments to engage in PPI research.<br>- The EPR would be likely to participate in PPI research again.<br>- Positive impacts of PPI on the research were the patient information sheet, participant manual, phone stand, newsletter, etc.<br>- No negative experiences of PPI on research were identified. |

### Policy and Practice

In the Policy and Practice section, both checklists reported that the design strategy and reason for PPI was clearly defined prior to the beginning of the trial. Both PPI member and researcher believed the goal of PPI was to improve the quality of research, and therefore, improve the quality of life of patients.

> *“The goal was to include PPI members as research partners throughout the entire research process. We wanted to do this in order to improve the quality, relevance, and acceptability of the research for people with MS” – Researcher form.*

> *“To improve the quality of the research, which would ultimately improve the quality of life of people living with MS, in particular their cognition.” – PPI member form*

In terms of resources, both agreed that sufficient time was given to learn about the project and for PPI members to provide input and deliberation in the process. Each form found that training materials were provided, however, the PPI form added that while they were given a manual, *“there was no training on how to perform my role”*(PPI member form).

The researcher and PPI member survey concluded that managing expenses was an issue for PPI members. They were compensated for their contribution by gift card, salary or one-off payment directly into their bank account. The researchers found difficulty with timely distribution of compensation (though this may have been more so an artefact of institutional finance procedures than a reflection on researcher motivations).

In total, six PPI members were involved and were recruited through engaging a voluntary organisation relevant to the trial’s focus (i.e. multiple sclerosis). While both teams found that this working environment created trust and respect for individual realities and mutual learning, the PPI team member was unsure if they initially felt prepared to work with the research team. There was mutual agreement that sufficient time for gaining familiarity with the trial and for input and deliberation was granted. The research team stated they identified health needs and catered for them by allowing accessible facilities for in-person meetings; however, the PPI member team found these facilities unsuitable for their needs.

> *“When meeting took place in person, we ensured parking was available nearby and accessible buildings were used. Meetings were held at a time that suited PPI members.” – Researcher form*

> *“The university found it very difficult to find a place for me to store a mobility scooter, and the eventual location was not suitable. The disabled toilets were not solely for disabled people, but for everyone, which defeats their purpose. The university did not provide me with equipment supports, which I had to provide from my own resources.” – PPI member form*

There were reciprocal views that the language used within intervention materials was translated into ‘plain English’ from academic language. The PPI checklist expressed that without experience or background in teamwork, PPI members may struggle to perform their role.

> *“I had experience in my previous work life of working in groups, and I also had experienced what research is like when I was doing my degree. This made the role easier for me, however, if I didn’t have this prior knowledge, it would have been more difficult to perform the role.” – PPI member form*

There was mutual agreement that the participants represented the target population; however, the research team did not make an extra effort to include underserved communities (e.g. given that underserved communities were not the target focus of the research, rather those with multiple sclerosis). Both surveys found that communication methods were adequate and allowed for honest conversations to support new ideas and decisions. The two forms also agreed that these methods were flexible as engagement was forced online due to the COVID-19 pandemic. PPI member involvement allowed contribution and implementation to decision-making. Table 5, below, summarises the main points from this theme from the perspective of the researcher and the PPI member.

### Participatory Culture

The second thematic section of the EPPIIC reported that PPI inclusion in this trial led to further collaborations between the research team and PPI members (e.g. in joint presentations and articles not directly related to the COB-MS). Reports were also provided on PPI members co-presenting results of the research and their perspectives to different audiences. One PPI member, to date, was cited as a co-author, while the PPI group have been acknowledged on multiple publications. Involvement in research facilitated the PPI members to gain knowledge on how PPI can influence the quality of research.

> *“I am more knowledgeable of the research process and know how the patient can help improve the quality of research.” – PPI member form*

Further results are presented from this section in Table 6.

### Influencing the Outcomes of PPI

The final section of the EPPIIC focused on the strengths of and challenges faced by PPI in the COB-MS trial. Specific examples included challenges in organisation of meetings when PPI were not responsible for chairing them, as well as wider organisational constraints such as the institution’s processes around payment. The PPI member reported challenges with the facilities of the institution but found that a clear sense of purpose was gained from their involvement in the study.

> *“I was able to use my experience of something negative, chronic illness, for something positive. It gave me a purpose again.” – PPI member form*

Conceptual developments that emerged concerned the creation of the role of ‘*Embedded Patient Researcher*’. This role was defined as it involved a greater depth of involvement in comparison to the PPI advisory panel. This was the first time this had been done in this institution. The research team also identified ‘thorough evaluation’ as an important consideration for future PPI inclusion.

## Discussion

The variety of Public and Patient Involvement typologies brings forth great challenge in the development of suitable evaluation tools, applicable across wide-ranging PPI scenarios. Many agree that no individual assessment method can be used (21), which is a barrier to comparing PPI between trials (27). However, it may be the case that there is a lack of checklists providing sufficient flexibility to assess different PPI contexts. As developed in the current research, the EPPIIC may have the necessary adaptability, as it provides space and opportunities for expression of PPI efforts, in broader contexts, that more specific questioning may miss. While the primary goal of such evaluation exists as an effort to improve PPI standards for future research, identifying tokenistic trials that include PPI just as part of an ‘integrated research agenda’ is also crucial (37). Although challenging, through consideration of comprehensiveness and attention to PPI specific questioning, it is possible to compare and contrast the strengths and weaknesses of various trials.

### Overall Interpretation of Results

Results from our research suggest that the use of open-ended responding at the end of each theme facilitated flexibility in reporting on topics not otherwise addressed.

Notably, a joint qualitative and quantitative approach responding/assessing is seldom used in other checklists but is a necessary component to include for purposes of ensuring a comprehensive evaluation. Qualitative research allows open questioning to assess opinions which allows broader understanding when compared to asking direct quantitative questions (39). Using open ended questions also allowed for individualised insight within concepts, a much-valued aid in further improvement and assessment, which helps to avoid the ‘tyranny of majority’, in which the generalised opinion dominates individual voices (40), particularly in specific contexts, like those that arise in research interventions This is important in encouraging diverse groups to participate in trials, as facilitating PPI means accommodating to each member’s reality, instead of allowing a needs assessment hospitable to the majority to cover all participants (25).

The EPPIIC, developed in the current research, includes process and outcome metrics, differing from some tools that only consider process-based evaluation (30). The themes included in the checklist identify the seemingly most vital areas of PPI in research (e.g. practice, culture, and outcomes). Importantly, this checklist can also be used when planning PPI activities – see Figure 1 below:

**Fig 1.**
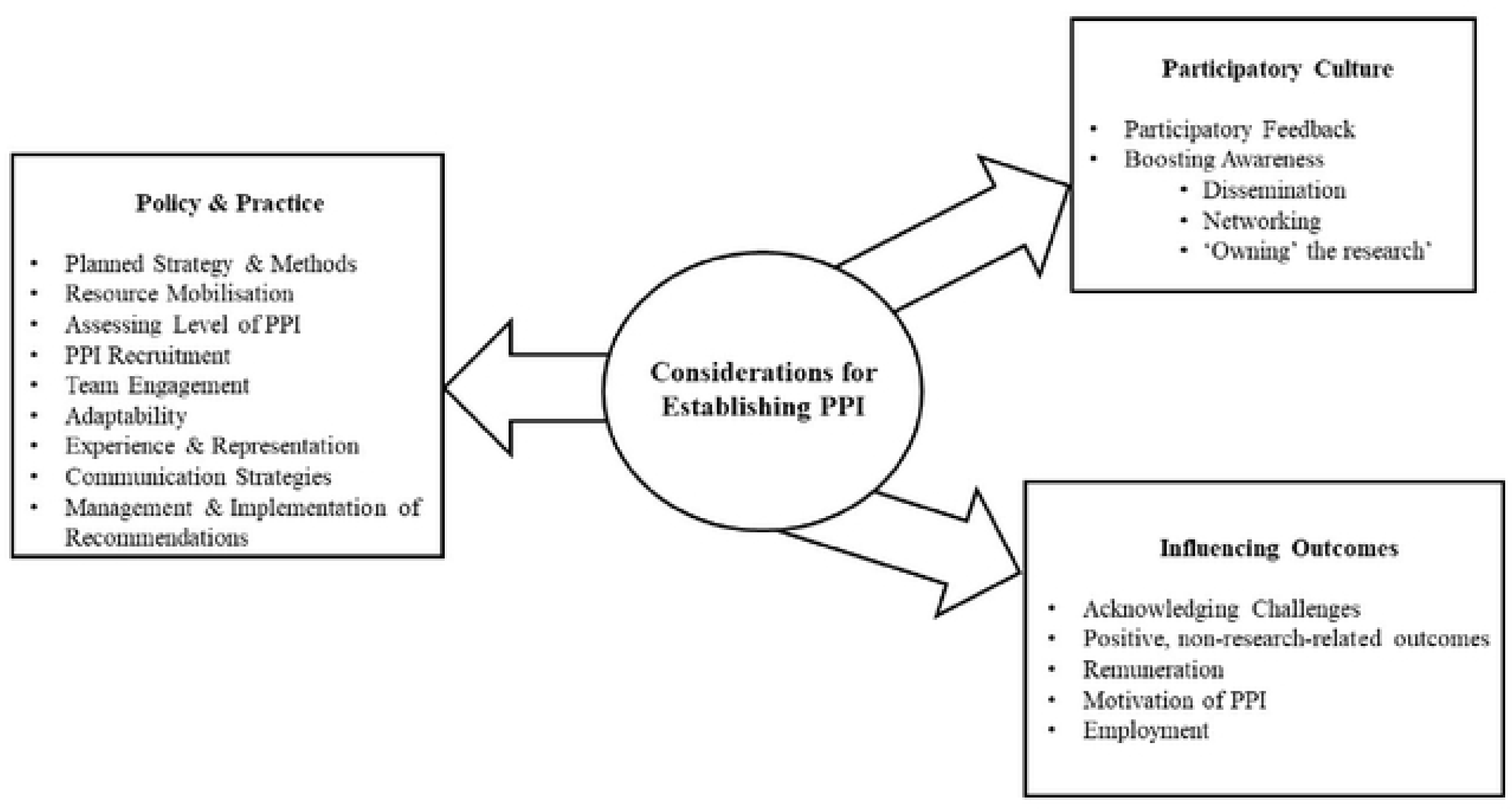
Considerations for Establishing PPI, adapted from the EPPIIC.

The largest section of the EPPIIC centres around the structural organisation of PPI within studies, a considerable area of downfall in previous PPI trials (32). These questions used within the framework pose as surrogates for understanding the factors that led to ‘good’ or ‘bad’ involvement experiences; for example, with respect to training or reimbursement (33). Some questioning requires an understanding of perspective; for example, participants agreeing that they would participate in the future reflects a positive outlook on their experience (28).

The themes were developed to thoroughly understand the levels of integration of PPI within trials. Often trials adapt the ‘one-off model’ meaning the research team has decided to include PPI for a specific reason, without considering the wider benefits of collaboration (35). It is often more transactional, with PPI members filling a consultant role rather than as a fellow researcher. Alternatively, the ‘fully intertwined model’ focuses on true participation and PPI member integration within the trial. It reflects the true purpose of PPI and adds the potential to gain all possible benefit, for both the research and the PPI team. Awareness of the approach of integration used reveals the level of consideration and preparation made to include PPI.

The use of two separated perspectives (researcher and PPI member) provided opportunity for divergent focuses which is a vital component of PPI, (28;40). Highlighting disparity in opinions between participants is the first step in improving future efforts and reinforces the efficacy of PPI methods. In order to provide their opinions, evaluators must be in an environment of honesty, openness, and respect (31). Providing separate EPPIICs to both researchers and PPI members facilitated this, as one form was not influenced by the opinions of others. In consideration of the PPI member’s checklist, it was important to adapt the checklist’s language to ensure accessibility and avoid unnecessarily technical jargon and terminology. Other than amendments to language and phrasing, the questions in each checklist remained largely a reflection of each other.

The application of the EPPIIC to the COB-MS feasibility trial provided insight into PPI factors that were beneficial and those that were challenging. The ‘Policy and Practice’ theme revealed that there was general agreement over the reasoning for including PPI in the research and the development of an environment of respect and trust. There was slight disparity within the suitability of facilities used and training resources mobilised. A good relationship between PPI members and researchers was revealed, with any conflict being found to be constructive and yielding an acceptable solution. There was a pre-planned structure to the inclusion and engagement of PPI within COB-MS, which other similar studies/partnerships have lacked, thus leading to subsequent deterioration of collaboration (41). A review assessing the impact of PPI within MS specific trials found that while many different approaches may be taken, inclusion in ‘trial design’ was crucial to giving PPI members power to impact the research and various communication channels are required to enhance participation, as utilised in this trial (42). Other consistent findings between the current research and Gray and colleagues’ review (42) were included the attitude of PPI members towards research, as well as the importance and methods of reimbursement. The benefits of PPI organisation and practice were reflected in the decisions that led to higher-quality research, as a direct result of PPI.

The theme of ‘Participatory culture’ showed a ‘fully intertwined’ partnership as PPI members were involved in citation and publication. The impact of PPI in this trial also led to further collaborations for the PPI members and researchers. The team sought feedback from the PPI team to allow them to express any concerns. This shows a true effort to involve the PPI team as fellow researchers and reflected a good relationship between the researcher and PPI member team.

The assessment of outcome-based metrics was included within the ‘Influencing the outcomes of PPI’ section. This again reflected that while accommodations were made for PPI involvement, the institution did not seem entirely suitable for PPI members to access. For example, a lack of equipment supports or facilities to store mobility aids. However, this was overcome, to some extent, by the adaptation of online meetings during the COVID-19 pandemic. This theme also exhibited insight into the effect of involvement for PPI members, as they found purpose in increasing the quality of research – feeling the ability to turn their negative experiences into positive input into possible clinical intervention. Positive impacts that PPI brought further included recruitment, proper engagement with participants, developing patient materials and communication and collaboration. Both parties agreed that they would get involved in PPI research in the future, reflecting a positive outlook on their own involvement.

### Limitations and Considerations for Future Research

Another step towards achieving an ideal PPI evaluation in future research would be the use of a possible weighted scoring system for the EPPIIC, where “positive” and “negative” PPI factors could be evaluated within each theme and metricised. An example of the ‘Rifkin Spider-gram’ (25) being applied to PPI evaluation using the EPPIIC is shown below in Figure 2.

**Fig 2:**
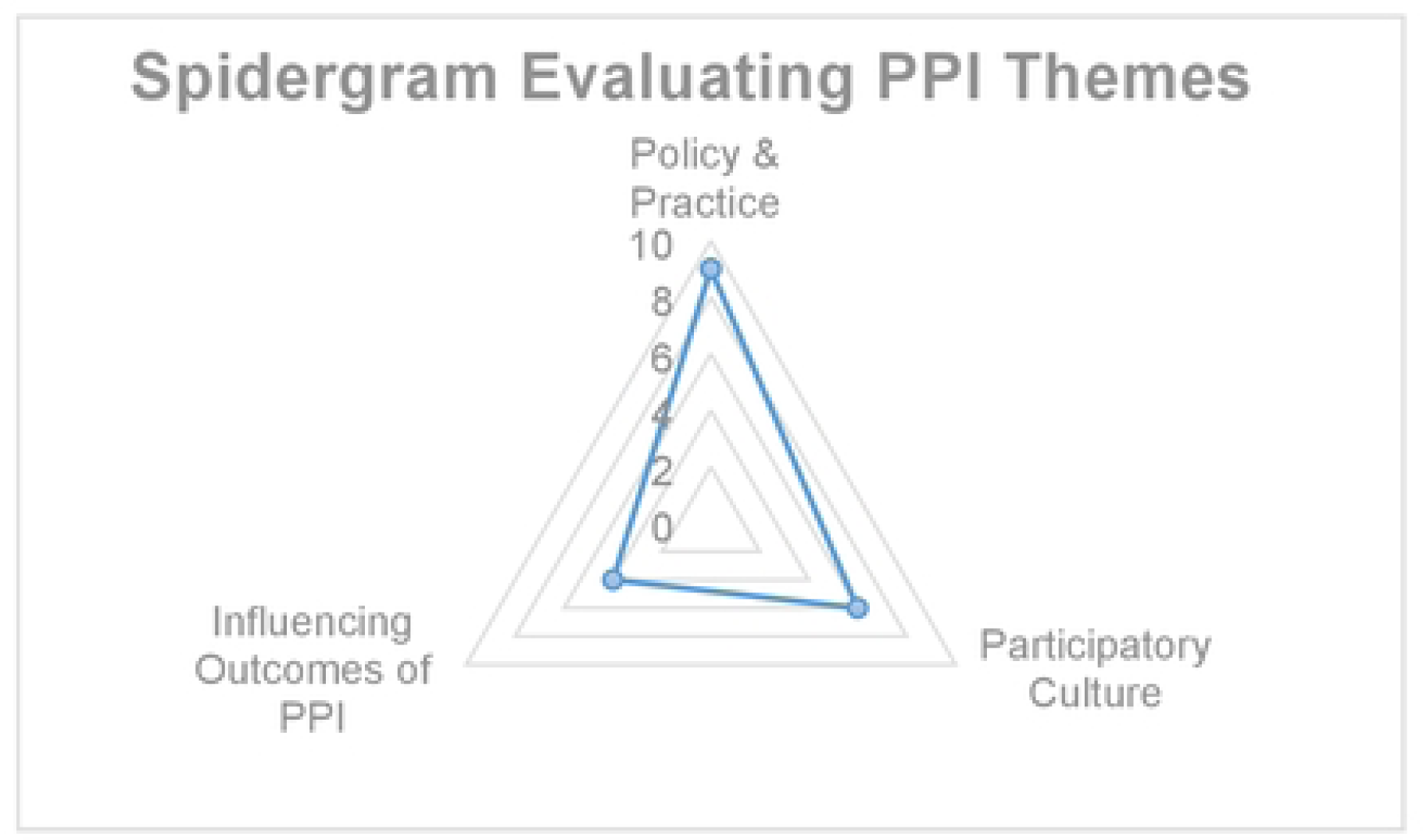
Spidergram displaying example of scoring system for PPI evaluation, using overarching themes as described previously. This example shows a score of 9 for Policy & Practice, 6 for Participatory Culture, and 4 for Influencing Outcomes of PPI.

However, as not all questions might receive the same weighting, as one factor could be deemed more crucial to PPI than another, a weighted system would have to be decided within a cohort of PPI members and researchers, which was not possible to carry out in this study.

Another challenge of implementing a scoring system is the decision of what would constitute ‘positive’ and ‘negative’ PPI impacts (13), as this is likely to vary considerably across trials/research settings. An example of such a scoring/weighting system is used by the STEPP guideline, in which PPI inspired changes to the study are valued as a positive impact.

However, in reality, non-implementation of such changes is not always a bad thing. The PPI that is shown in this instance is the ability of the research and PPI team to collaborate and discuss the issue and why a proposed solution is not possible, or maladaptive to the research. Therefore, the labelling of right and wrong needs to be carefully considered. It might also be suggested that such a weighting system might be arbitrary in the light of the potential richness that can be achieved through qualitative responding, as is the case for the current checklist.

Another area for future research would be the an evaluation of PPI throughout the trial, rather than just at the end (31). This feedback could guide PPI efforts to engage more efficient methods within the trial and make amendments, as necessary, in real time. Short assessments following meetings evaluating the effectiveness of communication and involvement aid the effort to improve future meetings and collaboration. Such an evaluation could also give all team members an opportunity to express any issues or queries anonymously, so that their concerns may be discussed at subsequent meetings. The EPPIIC was applied retrospectively to the COB-MS feasibility trial, however, in the future, an evaluation could be carried out alongside the research itself (e.g. once every three or six months, perhaps relative to the research’s life-cycle), presenting an opportunity to compare the final assessment to understand the impact of continuous evaluation.

### Conclusion

The EPPIIC, as developed in the current research, expressed a constructivist paradigm that focuses on reflection and notation for future improvement rather than a cynical criticism of possible shortcomings. Having both PPI and Researcher forms of the EPPIIC available allowed for flexibility to evaluate any intereventional study using PPI methods. It is hoped this tool leads to further improvement within PPI methods to facilitate use in increasing numbers of clinical trials and studies that will pave the way for optimal research and clinical developments in the years to come.

Overall, the inclusion of PPI within this trial was highly beneficial and was carried out well within the COB-MS team. There is evidence of the maintenance of a good relationship between parties, which is crucial to enabling proper (and future) involvement (26). The separate EPPIICs allowed for people to express their thoughts and opinions regarding components of PPI. By comparing perspectives, both strengths and weaknesses of PPI in this trial were highlighted. Adaptation and flexibility were exhibited throughout the trial, which appeared to lead to the achievement of high-quality PPI. Both parties independently expressed recommendations for future improvements, which is the ideal objective of PPI evaluation (37). The EPPIIC worked well to highlight the organisation of PPI and the culture created within the team.

The findings from this research suggests potential for the EPPIIC’s use in future research, as appropriate. We invite researchers and PPI members alike to use the checklist as a means of evaluating PPI within their own research.

## Data Availability

All relevant data are within the manuscript and its Supporting Information files.

## Acknowledgements

We wish to acknowledge the invaluable support and input from the COB-MS PPI group throughout this research and the associated COB-MS work.

## Supporting Information

**S1 Appendix. Evaluation of PPI for Interventional research Checklist (EPPIIC). EPPIIC (PPI Version)**

**S2 Appendix. Evaluation of PPI for Interventional research Checklist (EPPIIC). EPPIIC (Researcher Version)**

## References

1. Hewlett S, Wit M, Richards P, Quest E, Hughes R, Heiberg T, et al. Patients and professionals as research partners: challenges, practicalities, and benefits. Arthritis Rheum. 2006;55(4):676–80.

2. Pizzo E, Doyle C, Matthews R, Barlow J. Patient and public involvement: how much do we spend and what are the benefits? Health Expect. 2015;18(6):1918–26.

3. Jackson D, Moorley C. ‘Nothing about us without us’: embedding participation in peer review processes. J Adv Nurs. 2022;78(5):e75–e6.

4. Selman LE, Clement C, Douglas M, Douglas K, Taylor J, Metcalfe C, et al. Patient and public involvement in randomised clinical trials: a mixed-methods study of a clinical trials unit to identify good practice, barriers and facilitators. Trials. 2021;22(1):735.

5. Blackburn S, McLachlan S, Jowett S, Kinghorn P, Gill P, Higginbottom A, et al. The extent, quality and impact of patient and public involvement in primary care research: a mixed methods study. Res Involv Engagem. 2018;4:16.

6. Hoddinott P, Pollock A, O’Cathain A, Boyer I, Taylor J, MacDonald C, et al. How to incorporate patient and public perspectives into the design and conduct of research. F1000Res. 2018;7:752.

7. Joyce R, Dwyer CP & Hynes S. M. (2021). Twelve months into a feasibility trial: reflections on three experiences of public and patient involvement in research [version 2; peer review: 3 approved]. HRB Open Research, 4:11 10.12688/hrbopenres.13205.2

8. Tomlinson J, Medlinskiene K, Cheong VL, Khan S, Fylan B. Patient and public involvement in designing and conducting doctoral research: the whys and the hows. Research involvement and engagement. 2019 Dec;5:1–2.

9. Smits, D. W., Van Meeteren, K., Klem, M., Alsem, M., & Ketelaar, M. (2020). Designing a tool to support patient and public involvement in research projects: the Involvement Matrix. Research involvement and engagement, 6(1), 1–7.

10. Russell J, Fudge N, Greenhalgh T. The impact of public involvement in health research: what are we measuring? Why are we measuring it? Should we stop measuring it?. Research involvement and engagement. 2020 Dec;6:1–8.

11. Boivin A, L’Esperance A, Gauvin FP, Dumez V, Macaulay AC, Lehoux P, et al. Patient and public engagement in research and health system decision making: A systematic review of evaluation tools. Health Expect. 2018;21(6):1075–84.

12. Greenhalgh T, Hinton L, Finlay T, Macfarlane A, Fahy N, Clyde B, et al. Frameworks for supporting patient and public involvement in research: Systematic review and co-design pilot. Health Expect. 2019;22(4):785–801.

13. Rowe G, Frewer, L. J. Public Participation Methods: A Framework for Evaluation. Science, Technology, & Human Values. 2000;25(1):3–29.

14. Dwyer, C.P., Alvarez-Iglesias, A., Joyce, R., Counihan, T. J., Casey, D. & Hynes, S.M. (2020). Evaluating the feasibility and preliminary efficacy of a Cognitive Occupation-Based programme for people with Multiple Sclerosis (COB-MS): protocol for a feasibility cluster-randomised controlled trial. Trials, 21(1), 269. 10.1186/s13063-020-4179-5

15. Dwyer, C.P., Alvarez-Iglesias, A., Joyce, R. Counihan, T. J., Casey, D. & Hynes, S.M. (2023). Evaluating the feasibility and preliminary efficacy of a Cognitive Occupation-Based programme for people with Multiple Sclerosis (COB-MS): an update to the protocol for a feasibility cluster-randomised controlled trial. Trials 24, 48. 10.1186/s13063-023-07080-y

16. Cooke, A., Smith, D., & Booth, A. (2012). Beyond PICO: the SPIDER tool for qualitative evidence synthesis. Qualitative health research, 22(10), 1435–1443.

17. Braun V, Clarke V. One size fits all? What counts as quality practice in (reflexive) thematic analysis?. Qualitative research in psychology. 2021 Jul 3;18(3):328–52.

18. Byrne D. A worked example of Braun and Clarke’s approach to reflexive thematic analysis. Quality & quantity. 2022 Jun;56(3):1391–412.

19. Garratt A, Sagen J, Borosund E, Varsi C, Kjeken I, Dagfinrud H, et al. The Public and Patient Engagement Evaluation Tool: forward-backwards translation and cultural adaption to Norwegian. BMC Musculoskelet Disord. 2022;23(1):556.

20. Public and Patient Engagement Collaborative MU. Public and Patient Engagement Evaluation Tool (PPEET). 2018.

21. Group PS. The Public Involvement Impact Assessment Framework: Executive Summary. Lancaster University; 2014.

22. System HPOIH. Engagement Toolkit. 2016.

23. Maybee A, Clark, B., McKinnon, A., Angl, E. N. Patients as Partners in Research: Patient/Caregiver Surveys Patients Canada; 2016.

24. Maybee A, Clark, B., McKinnon, A., Angl, E. N. Patients as Partners in Research: Researcher Surveys Patients Canada; 2016.

25. Rifkin SB, Muller F, Bichmann W. Primary health care: on measuring participation. Soc Sci Med. 1988;26(9):931–40.

26. Committee NCE. Framework for Public Involvement in Clinical Effectiveness Processes. 2018.

27. South J, Fairfax P, Green E. Developing an assessment tool for evaluating community involvement. Health Expect. 2005;8(1):64–73.

28. Dukhanin V, Topazian R, DeCamp M. Metrics and Evaluation Tools for Patient Engagement in Healthcare Organization-and System-Level Decision-Making: A Systematic Review. Int J Health Policy Manag. 2018;7(10):889–903.

29. Foley L, Kiely B, Croke A, Larkin J, Smith SM, Clyne B, et al. A protocol for the evaluation of the process and impact of embedding formal and experiential Public and Patient Involvement training in a structured PhD programme. J Multimorb Comorb. 2021;11:26335565211024793.

30. Wilton P, Neville D, Audas R, Brown H, Chafe R. An Evaluation of In-Person and Online Engagement in Central Newfoundland. Healthc Policy. 2015;11(2):72–85.

31. Arora PG, Krumholz LS, Guerra T, Leff SS. Measuring Community-Based Participatory Research Partnerships: The Initial Development of an Assessment Instrument. Prog Community Health Partnersh. 2015;9(4):549–60.

32. Woodland RH, Hutton, M. S. Evaluating Organisational Collaborations: Suggested Entry Points and Strategies. American Journal of Evaluation. 2012;33(3):366–83.

33. Staniszewska S, Brett J, Simera I, Seers K, Mockford C, Goodlad S, et al. GRIPP2 reporting checklists: tools to improve reporting of patient and public involvement in research. Res Involv Engagem. 2017;3:13.

34. Council SH1. The Participation Toolkit: Supporting Patient Focus and Public Involvement in NHS Scotland 2014.

35. Wilson P, Mathie E, Keenan J, McNeilly E, Goodman C, Howe A, et al. ReseArch with Patient and Public invOlvement: a RealisT evaluation - the RAPPORT study. Health Services and Delivery Research. Southampton (UK)2015.

36. Broerse JE, Zweekhorst MB, van Rensen AJ, de Haan MJ. Involving burn survivors in agenda setting on burn research: an added value? Burns. 2010;36(2):217–31.

37. Abma TA, Broerse JE. Patient participation as dialogue: setting research agendas. Health Expect. 2010;13(2):160–73.

38. Kreindler SA, Struthers A. Assessing the organizational impact of patient involvement: a first STEPP. Int J Health Care Qual Assur. 2016;29(4):441–53.

39. Cleland JA. The qualitative orientation in medical education research. Korean J Med Educ. 2017;29(2):61–71.

40. Tritter JQ, McCallum A. The snakes and ladders of user involvement: Moving beyond Arnstein. Health Policy. 2006;76(2):156–68.

41. Hughes J, Weiss J. Simple rules for making alliances work. Harv Bus Rev. 2007;85(11):122–6, 8, 30-1 passim.

42. Gray E, Amjad A, Robertson J, Beveridge J, Scott S, Peryer G, et al. Enhancing involvement of people with multiple sclerosis in clinical trial design. Mult Scler. 2023;29(9):1162–73.

